# Clinical characteristics and outcomes of patients with COVID-19 and ARDS admitted to a third level health institution in Mexico City

**DOI:** 10.1101/2020.09.12.20193409

**Authors:** Carmen M. Hernández Cárdenas, Carlos Torruco Sotelo, Felipe Jurado, Héctor Serna-Secundino, Cristina Aguilar, José G. García-Olazarán, Diana Hernández García, Gustavo Lugo Goytia

## Abstract

**Background:** In December 2019, the first cases of severe pneumonia associated with a new coronavirus were reported in Wuhan, China. Severe respiratory failure requiring intensive care was reported in up to 5% of cases. There is, however, limited information available in Mexico.

**Objectives:** The purpose of this study was to describe the clinical manifestations, and outcomes in a COVID-19 cohort attended to from March to May 2020 in our RICU. In addition, we explored the association of clinical variables with mortality.

**Methods:** The first consecutive patients admitted to the RICU from March 3, 2020, to Jun 24, 2020, with confirmed COVID-19 were investigated. Clinical and laboratory data were obtained. Odds ratios (ORs) were calculated using a logistic regression model. The survival endpoint was mortality at discharge from the RICU.

**Results:** Data from 68 consecutive patients were analyzed. Thirty-eight patients survived, and 30 died (mortality: 44.1 %). Of the 16 predictive variables analyzed, only 6 remained significant in the multivariate analysis [OR (95% confidence interval)]: no acute kidney injury (AKI)/AKI 1: [.61 (.001;.192)]; delta lymphocyte count: [.061 (.006;.619)]; delta ventilatory ratio: [8.19 (1.40;47.8)]; norepinephrine support at admission: [34.3 (2.1;550)]; body mass index: [1.41 (1.09;1.83)]; and bacterial coinfection: [18.5 (1.4;232)].

**Conclusions:** We report the characteristics and outcome of patients with ARDS and COVID-19. We found six independent factors associated with the mortality risk: delta lymphocyte count, delta ventilatory ratio, BMI, norepinephrine support, no AKI/AKI 1, and bacterial coinfection.

## Introduction

In December 2019, the first cases of severe pneumonia associated with a new coronavirus were reported in Wuhan, China. Severe acute respiratory syndrome coronavirus 2 (SARS-CoV-2) has spread rapidly to other countries, causing a pandemic. It binds to the cellular receptors of angiotensin-converting enzyme 2 to penetrate the cells^1^. Severe pulmonary inflammation with exudative diffuse alveolar damage and massive capillary congestion often accompanied by microthrombi^2^, which translates physiologically in ventilation-perfusion inequalities, leads to severe acute hypoxemic respiratory failure, requiring intensive care and mechanical ventilation in up to 5% of cases^3^.

The experience with COVID-19 in China, Europe, and the United States has been widely reported^4-8^, however, there is limited information available in Mexico and other Latin American countries. The Instituto Nacional de Enfermedades Respiratorias s (INER), Ismael Cosío Villegas was declared as a COVID-19 center, and in the beginning of March, the respiratory intensive care unit (RICU) received the first patients with acute hypoxemic respiratory failure secondary to SARS-CoV-2 infection. The purpose of this study was to describe the clinical manifestations, laboratory data, imaging characteristics, and outcomes in a COVID-19 cohort attended from March to June 2020 in our RICU. In addition, we explored the association of clinical, laboratory, and physiologic variables with mortality.

## Methods

### Patients and setting

The first consecutive patients admitted to the RICU from March 3, 2020 to June 24, 2020 with confirmed SARS-CoV-2 infection (positive polymerase chain reaction test result from a nasopharyngeal sample or tracheal aspirate) were investigated. The INER is a tertiary-level teaching and research hospital that serves the open population of Mexico City and states of the Republic. The studies were granted exemption by the hospital institutional review boards. The requirement for informed consent was also waived.

### Data collection and definitions

Clinical and laboratory data were obtained upon admission and daily during the patients’ stay in the unit. High-resolution computed tomography was performed in all patients prior to admission to the RICU. The information obtained was stored in an electronic database.

Acute respiratory distress syndrome (ARDS) was diagnosed in accordance with the Berlin definition^9^ and septic shock in accordance with the 2016 Third International Consensus Definition^10^. Acute kidney injury (AKI) was diagnosed in accordance with the Kidney Disease: Improving Global Outcomes (KDIGO) clinical practice guidelines^11^. Body mass index (BMI) was calculated as follows: weight (kg)/height (m)^2^. Bacterial coinfection was defined as a positive culture and compatible clinical picture. In cases where the cultures were negative, it was considered on the basis of the presence of persistent fever for more than 48 hours, leukocytosis, neutrophilia, increased procalcitonin level, and hemodynamic instability. The respiratory parameters [i.e., static compliance, driving pressure, and ventilatory ratio (VR)] were calculated using the formulas described in the table 2. The survival endpoint was mortality at discharge from the RICU.

### Statistical analysis

We used descriptive statistics to summarize the clinical data; the results were reported as medians and ranges. Meanwhile, the categorical variables were reported as frequencies. Odds ratios (ORs) were calculated using a logistic regression model. The models were fitted with univariate and multivariate inputs using the potential risk factors for COVID-19 mortality: age, BMI, AKI, absolute lymphocyte count, PaO_2_/FiO_2_ ratio, prone ventilation, static compliance, driving pressure, VR, use of vasopressors, and bacterial coinfection. SPSS version 21 (IBM Statistics, Armonk, New York) was used to analyze the data. Two-sided p-values of <.05 were considered statistically significant.

## Results

### Univariate analysis of mortality association

#### Clinical characteristics and laboratory results

Data from 68 consecutive patients admitted to the RICU from March 4, 2020, to June 24, 2020, were analyzed. Thirty-eight patients survived, and thirty died (mortality: 44.1%). Age, sex, comorbidity, and the clinical symptoms were not associated with mortality (Table 1). Conversely, the BMI was associated with an increased risk of mortality [OR, 1.10 (95% confidence interval {CI}, 1.001–1.21, p= .046)].

**Table 1.** Characteristics of the patients at to the RICU admission.

| **Patients characteristics** | **All patients (68)** |
| --- | --- |
| Age (years) | 50(23-73) |
| Sex (M/F) | 43/25 |
| BMI (Kg/m<sup>2</sup>) | 32.1(23.1-63.6) |
| Onset of symptoms to hospital admission (days) | 7(3-14) |
| **Comorbidities** | **No(%) of patients** |
| Diabetes | 16(23.5) |
| Obesity | 34(50) |
| Systemic arterial hypertension | 13(19.1) |
| **Admission symptoms** | **No(%) of patients** |
| Fever | 64(94.1) |
| Cough | 68(100) |
| Shortness of breath | 68(100) |
| Muscle pain | 54(79.4) |
| Headache | 60(88.2) |
| Diarrhea | 12(17.6) |
| **CT scan findings** | **No(%) of patients** |
| Peripheral bilateral ground glass opacities | 68(100) |
| Crazing paving pattern | 26(38.2) |
| Focal consolidation | 46(67.6) |
| **Laboratory measures** | **Mean(range)** |
| White blood cell count/μl | 10.11(5-24.4) |
| Lymphocyte count / μl | 829(100-2600) |
| Hemoglobin (gr/dl) | 14.9(10.2-18.4) |
| Platelet count $\times 10^3$ /μl | 213(37-613) |
| Creatinine (0.7-1.2 mg/dl) | 1.42(0.58-5.04) |
| Total bilirubin (0.3-1.2 mg/dl) | 0.86(0.25-2.35) |
| Aspartate aminotransferase(17-63 U/L) | 89(22-467) |
| Alanine aminotransferase(15-41 U/L) | 62(18-215) |
| Lactic dehydrogenase (98-192 UI/L) | 487(208-845) |
| Glucose (74-118 mg/dl) | 156(84-334) |
| Creatinine kinase (49-397 UI/L) | 952(29-7340) |

From the laboratory tests performed, no associations with mortality were found for the following variables: lactic dehydrogenase level, creatinine level, hepatic test result, troponin level, fibrinogen level, D-dimer level, total leukocyte count, total neutrophil count, total lymphocyte count, hemoglobin level, and platelet count. The only variable that showed a significant association with mortality was the Δ lymphocyte count [OR, .99 (95% CI, .99–1.0); p=.014], defined as the difference between the total admission lymphocyte count and the total lymphocyte count in the last evaluation before discharge from the RICU. Based on this result, an increase in the total lymphocyte count showed a protective effect on mortality.

#### Cardiovascular and respiratory variables

The systolic, diastolic, and mean pressures was not associated with mortality. However, norepinephrine support was associated with a significant increase in the mortality risk [OR, 3.4 (95% CI, 1.1–10.4); p=.031)].

All patients received invasive protective mechanical ventilation with tidal volumes of 4–8 mL/kg of predicted body weight. The monitored respiratory parameters (i.e., static compliance, PEEP, driving pressure, and VR at admission) were not associated with mortality. Although the VR at admission showed no association with mortality, the difference between the VR determined at admission and the VR determined on the last day of invasive mechanical ventilation showed a significant association [OR, 4.7 (95% CI, 1.8–11.9); p=.001)]. The PaO_2_/FiO_2_ ratio showed a significant protective effect for mortality (higher values were protective) [OR, .98 (95% CI, .97–.99); p=.037]. Ventilation in the prone position did not show a protective effect [OR, .38 (95% CI, 0.12–1.10); p=.07)].

### Multivariate analysis of mortality association

The variables that showed a significant association with mortality in the univariate analysis were included in the multivariate analysis. Linearity of the continuous variables with respect to the logit of the dependent variable was assessed via the Box-Tidwell procedure^12^. Bonferroni correction was applied using all six terms in the model, resulting in statistical significance being accepted at p-values of <.0083^13^. Based on this assessment, all continuous independent variables were found to be linearly related to the logit of the dependent variable. The analysis of the standardized residuals did not show outliers (greater than ±2 SD). The logistic regression model revealed significant findings [χ^2^ (4) =48.023, p<.0005]. The model explained 75.1% (Nagelkerke R^2^) of the variance in mortality and correctly classified 89.7% of the cases. The sensitivity was 76.7%; specificity, 78.6%; positive predictive value, 75.8%; and negative predictive value, 79.3%. Of the 16 predictive variables, only 6 remained statistically significant: no AKI/AKI 1, Δ lymphocyte count, Δ VR, norepinephrine support at admission, BMI, and bacterial coinfection (as shown in Table 2). An increasing Δ VR was associated with an increased risk of mortality; however, an increasing Δ absolute lymphocyte count was associated with a reduction in the risk of mortality. No AKI/AKI 1 showed a strong protective effect [OR, .019 (95% CI, .001–.400)]; norepinephrine support at admission and bacterial coinfection were also associated with a high risk of mortality [OR, 34.3 (95% CI, 2.1–550]; p=.012 and OR, 18.5 (95% CI, 1.4–232); p=.023, respectively]. The area under the ROC curve was .876 (95% CI, .790–.962), which was an excellent level of discrimination according to Hosmer et al.^14^

**Table 2.** Univariate and multivariate logistic regression analysis of risk factors associated with mortality.

| Variable | Univariate<br>OR | 95%C.I | P value | Multivariate<br>OR | 95%C.I | P value |
| --- | --- | --- | --- | --- | --- | --- |
| Age (years) | 1.024 | .980-1.070 | .296 |  |  |  |
| BMI (Kg/m<sup>2</sup>) | 1.101 | 1.001-1.211 | .048 | 1.41 | 1.09-1.83 | .009 |
| DHL (UI/L) | 1.001 | .999-1.003 | .212 |  |  |  |
| Creatinine (mg/dl) | .997 | .689-1.441 | .986 |  |  |  |
| Lymphocytes (count /<br>μ) | 1.000 | .999-1.001 | .443 |  |  |  |
| Δ Lymphocytes(count /<br>μ | .999 | .998-1.000 | .014 | .061 | .006-.619 | .018 |
| PaO<sub>2</sub>/FiO<sub>2</sub> (mmHg) | .987 | .975-.999 | .037 |  |  |  |
| PaCO<sub>2</sub> (mmHg) | .983 | .935-1033 | .499 |  |  |  |
| Static compliance<br>(ml/cm H<sub>2</sub>O)§ | .993 | .941-1.047 | .796 |  |  |  |
| Driving pressure (cm<br>H<sub>2</sub>O)¶ | 1.108 | .923-1.33 | .271 |  |  |  |
| Ventilatory ratio & | .886 | .405-1.94 | .763 |  |  |  |
| Prono ventilation<br>(yes/no) | 2.669 | .924-7.716 | .070 |  |  |  |
| Δ Ventilatory Ratio | 4.76 | 1.89-11.98 | .001 | 8.19 | 1.40-47.8 | .019 |
| Norepinephrine<br>(yes/no) | 3.429 | 1.12-10.47 | .031 | 34.3 | 2.1-550 | .012 |
| Bacterial coinfection<br>(yes/no) | 2.318 | .809-6.64 | .118 | 18.5 | 1.4-232 | .023 |
| No AKI/AKI 1(yes/no) | .0420 | .005-.353 | .003 | .061 | .001-.192 | .006 |

§Respiratory-system compliance was calculated as the ratio of the tidal volume to the difference between inspiratory plateau pressure and PEEP
¶Driving pressure was calculated as the difference of the plateau pressure and PEEP
&Ventilatory ratio (VR), was calculated as [minute ventilation (mL/min) × PaCO<sub>2</sub> (mmHg)] /
[predicted body weight × 100 (mL/min) × 37.5 (mmHg)]

## Discussion

In this study, we described the characteristics of a cohort of critically ill patients with COVID-19 and ARDS and analyzed the factors associated independently with mortality. Our analyses showed that the demographic, clinical, radiological, and biochemical characteristics are similar to those reported previously by other groups in China, Europe, and the United States^4-8^. Six variables showed an independent association with mortality: Δ lymphocyte count, Δ VR, no AKI/AKI 1, norepinephrine use, BMI, and bacterial coinfection.

Lymphocytopenia has been described as a marker of the severity of SARS-CoV-2 infection and a predictor of mortality since the first reports in Wuhan, China^5,15^. In our patients, the mean lymphocyte count was low in the survivors and non-survivors on admission to the RICU; however, the difference did not show a significant association with mortality. Nevertheless, the Δ lymphocyte count (difference between the total lymphocyte count at admission and total lymphocyte count at discharge) showed a strong protective effect on mortality. Thus, an increase was associated with a decrease in the risk of mortality, while a decrease was associated with a significant increase in the risk. This result is consistent with the rapid and dramatic restoration of peripheral T lymphocytes in patients who recovered from SARS, as reported by Li et al.^19^. A recent meta-analysis supports lymphocytopenia on admission as a marker of severity and as a predictor of mortality in patients with COVID-19 (17). However, the results of this study should be interpreted considering that the study only analyzed reported studies of patients in China, used information from studies without a peer review, and had the composite outcome that considered not only mortality. Furthermore, a zero time is difficult to determine because patients are expected to differ in the stage of the inflammatory response caused by the infection. Based on our results, lymphocytopenia on admission may not be a reliable predictor of mortality and thus should be used with caution when making decisions. The Δ lymphocyte count seems to be a better predictor of mortality and could be useful for stratifying patients when analyzing clinical trial results or as a surrogate biomarker of therapeutic efficacy. The mechanism of lymphocyte reduction in COVID-19 remains unclear; however, several hypotheses have been advanced to explain its pathogenesis^18,19^.

Lymphopenia due to SARS-CoV-2 infection is the expression of a dysregulated immune response with a predominance of immunosuppression and is associated with a high risk of bacterial coinfection, septic shock, organ dysfunction, and mortality^20^. A dramatic reduction in the total lymphocyte count and CD8 and CD4 T cell subpopulations to levels similar to those seen in patients with severe AIDS has been reported in patients with COVID-19 ^21^. Recovery of the adaptive immune system with an increase in the number of T lymphocytes is necessary for the elimination of the virus. In this context, the use of steroids can have both favorable and unfavorable consequences. With an excessive inflammatory response, steroids may reduce organ damage and may be of benefit in the early stage. However, in patients in whom the immunosuppression response predominates, steroids would accentuate this response, increasing the risk of sepsis and mortality; the application of treatments that stimulate the immune system could be useful in these patients^22^. In this context, changes in the lymphocyte count could be used as a parameter for decision-making.

The VR is governed by the production of carbon dioxide (VCO2) and the ventilatory efficiency [1-(Vd/Ve)] and can be easily calculated at the patient’s bedside using ventilation and blood gas parameters. It correlates with the percentage of dead space and is also associated with an increased risk of mortality^23^. Previous studies on patients with ARDS and COVID-19 have reported a significant association between the VR at admission and mortality^24^. In our patients, we did not observe this association upon admission to the RICU. This disagreement with previous findings, which indicate a significant association at admission, may be related to the different zero times in the evolution of the cohorts, as it is difficult to accurately determine the start time of COVID-19 pulmonary lesions. However, the Δ VR showed a strong association with mortality in our cohort. Thus, a progressive increase in the VR could be considered as a marker of the mortality risk in patients with ARDS due to COVID-19. This result is consistent with the increase in the fraction of dead space in the first weeks of ARDS as an independent predictor of mortality, as reported by several studies^23,25,26^.

In summary, worsening of the VR in our cohort was independently associated with an increased risk of mortality. Similar to the tidal volume adjusted for the predicted weight, plateau pressure, driving pressure, and PaO_2_/FiO_2_ ratio, the VR should be monitored on a daily basis and used to make adjustments to the ventilatory parameters, always taking into account the aforementioned variables.

Although the PaO_2_/FiO_2_ ratio showed a significant association with mortality in the univariate analysis, the association was not maintained in the multivariate analysis; thus, it did not represent an independent effect on mortality in our cohort. The PaO_2_/FiO_2_ ratio is a key variable for the definition and determination of the severity of ARDS according to the Berlin definition^9^. However, in patients with COVID-19, the PaO_2_/FiO_2_ ratio has not been shown to be a strong predictor of mortality, which may be related to the heterogeneity described for ARDS due to COVID-19^27^. The majority of our patients showed a ground-glass image on tomography, without extensive consolidation images, as observable in most patients with ARDS not due to COVID-19. The main mechanism of hypoxemia in these patients in the initial phase is an abnormality in the distribution between ventilation and blood flow; the latter is suggested to be due to abnormal vasoregulation^27^. This explains the rapid response of many patients to oxygen administration and the poor response at this stage to recruitment maneuvers because there are no extensive recruitable consolidation areas. Therefore, the PaO_2_/FiO_2_ ratio is not a physiological biomarker of the amount of pulmonary shunt and lung parenchymal damage in the early phase of ARDS due to COVID-19, which may explain why the PaO_2_/FiO_2_ ratio at this stage was not an independent predictor of mortality.

A high incidence of AKI and associated mortality has been reported in patients with COVID-19. Hirsch et al. reported an incidence of 36.6% in a cohort of 5449 patients with COVID-19 and a mortality of 35%. However, in patients with respiratory failure who required invasive mechanical ventilation, the incidence of AKI was 89.7%; in those who required hemodialysis, the mortality was 55%^28^. In our cohort, 74.1% of the patients developed some stage of AKI. Of them, 65.1% developed AKI 1, and only 25% did not present AKI during their evolution; meanwhile, 34% of the patients developed AKI 2 or AKI 3. The multivariate analysis showed that not developing AKI or AKI 1 had a strong protective effect on mortality. Thus, the application of preventive or therapeutic measures (implementation of the KDIGO supportive care guidelines)^11^ to avoid AKI or to prevent progression to more advanced stages in patients with AKI 1 should be a priority in critically ill patients with COVID-19.

Obesity has spread worldwide and is considered a pandemic with high morbidity and mortality. In Mexico, the prevalence of overweightness has been estimated at 71.2% in the adult population and that of obesity, defined as a BMI of >30 kg/m^2^, at 32.4%^29^. In our cohort, obesity showed a significant independent association with mortality. However, in patients with ARDS, a paradox has been reported, consisting of lower mortality in populations with obesity than in those without obesity^30^. Our data on COVID-19 contradict this paradox, and there are scientific arguments to support the pathogenic role of obesity in the severity of SARS-CoV-2 infection: 1. Adipocytes activate the inflammatory cascade, which increases the risk of thromboembolism and susceptibility to the cytokine storm^31^. 2. Obesity negatively alters respiratory mechanics^32^. 3. Alterations in mitochondrial bioenergetics in alveolar epithelial cells have been described in experimental models^33^, which could increase susceptibility to lung damage. 4. There is an experimental evidence of a compromise in innate and adaptive immunities owing to obesity^34^. In this context, obesity is a public health problem that may be associated with mortality during this pandemic.

### Study limitations

One limitation of the study is the small size of the cohort, which originated from a single third-level center in Mexico; the results may not then be extrapolated with certainty to other non-similar populations. However, our results are generally consistent with those reported in other larger cohorts by researchers in China, Europe, and North America.

Although the statistical logistic regression model showed a good discrimination performance, it should not be used for prediction purposes, as it requires internal and external evaluations as well as a larger number of patients. However, the association of the independent variables with the risk of mortality may help in better understanding the pathophysiological response of the organism to infection by SARS-CoV-2. In subsequent studies, these variables can be considered as potential predictors in the development of prognostic models.

Some of the strengths of the study are that it investigated a prospective cohort with a well-defined zero time for each patient and that the data were obtained in real time.

In summary, we described the clinical, laboratory, and radiological characteristics in a prospective cohort of critically ill patients with COVID-19 and the independent factors associated with mortality. Based on this information, it is possible to suggest some management recommendations in patients with COVID-19 who require intensive care; respiratory management based on low tidal volumes and adjustment of parameters according to the VR, measures to protect kidney function, adjustment of fluid balance according to volume responsiveness, avoidance of the use of immunosuppressants in patients who do not show lymphocyte recovery, strict measures to prevent nosocomial infections, and early detection and aggressive treatment when infection is suspected, and consideration of empirical antifungal therapy in patients with persistent lymphopenia and fever are simple measures that can potentially reduce the risk of mortality until an effective treatment against SARS-CoV-2 infection is available.

## Data Availability

data are available under request to the author

https://drive.google.com/file/d/1ShZifyMwgC36Hal_X7mBcHdTVpPNJVVH/view?usp=sharing

## Author contributions

All authors contributed to the data acquisition, patient care, and manuscript development.

